# Detection of SARS-CoV2 antigen in human saliva may be a reliable tool for large scale screening

**DOI:** 10.1101/2020.12.17.20248437

**Authors:** Priya Kannian, Chandra Lavanya, Krittika Ravichandran, Bagavad Gita Jayaraman, Pasuvaraj Mahanathi, Veeraraghavan Ashwini, Nagalingeswaran Kumarasamy, Gunaseelan Rajan, Kannan Ranganathan, Stephen J. Challacombe, Jennifer Webster-Cyriaque, Newell W. Johnson

## Abstract

**Introduction:** SARS-CoV2, the aetiological agent of the current COVID-19 pandemic, has been detected in saliva and recently implicated in several oral diseases. Collection of nasopharyngeal swabs (NPS) and detection by reverse transcriptase-polymerase chain reaction (RT-PCR) requires medical / technical expertise. A reliable and easy to handle point-of-care (POC) test is highly desirable, especially to curb transmission. Therefore, in this study, we evaluated a commercially available POC rapid antigen test (RAT) for the detection of SARS-CoV2 antigens in the saliva of RT-PCR confirmed positive and negative patients.

**Methods:** Thirty saliva samples of 10 saliva RT-PCR negative and 20 saliva RT-PCR positive patients were tested by RAT.

**Results:** RAT was negative in 10/10 (100%) RT-PCR-negative samples; positive in 9/20 (45%) RT-PCR-positive samples; concordance was 63% (p=0.001). Patients with positive RAT had higher virus copies in their NPS samples compared to the RAT-negative patients. This difference was also statistically significant (p=0.01).

**Conclusion:** Thus, the POC RAT may be used to detect SARS-CoV2 as a reliable tool for self-testing, large-scale population screening and emergency medical/dental screening. Patients negative by RAT should be confirmed by RT-PCR.

## Text

COVID-19, the current pandemic, is caused by SARS-CoV2, a respiratory RNA virus. Respiratory viruses are transmitted through secretions from the respiratory tract that are dispersed within close contacts. SARS-CoV2 primarily infects the epithelial and endothelial cells lining the respiratory mucosae and the ducts of minor mucous glands. A number of different respiratory tract samples, including nasopharyngeal swab (NPS), sputum, bronchial aspirate and bronchoalveolar lavage have been tested for the detection of SARS-CoV2 RNA [1]. The NPS rapidly became the favored sample, as it retrieves cells from a site where the virus perpetuates with minimal risk of transmission through aerosols [2]. However, collection of NPS is somewhat invasive and causes discomfort; it requires medical/technical expertise, which might not be available in remote villages, especially in developing countries like India. On the other hand, epithelial cells of the oral mucosa and particularly the ducts of the minor salivary glands, have been shown to carry large numbers of angiotensin converting enzyme-2 (ACE-2) receptors that bind the SARS-CoV2 virus [3,4]. Symptoms such as loss of taste and smell, xerostomia and oral ulcerations have been increasingly reported with COVID-19 pointing to salivary transmission of the SARS-CoV2 virus [5]. Further there is increasing evidence to date pointing to the involvement of the salivary gland by itself in COVID-19 with acute parotitis being commonly reported [6].

Saliva is now accepted as an ideal diagnostic fluid for numerous diseases, including cardiovascular diseases and several cancers, with detectable levels of antibodies, antigens, DNA, RNA and microorganisms, with sensitivities and specificities comparable to serum [8]. This combined with its non-invasive method of self-collection, which does not require technical expertise and has less risk of disease transmission, makes saliva an attractive choice as a diagnostic specimen [9,10]. Early and quick detection of SARS-CoV2 is of prime importance in containing its spread. Most rapid antigen kits in commercial use are based on NPS specimens. In this study we evaluated the utility of a SARS-CoV2 antigen kit using drooled saliva samples from laboratory-confirmed SARS-CoV2 RT-PCR positive patients.

The study was approved by the VHS-Institutional Ethics Committee (proposal #: VHS-IEC/69-2020). Saliva samples from 30 patients previously tested for SARS-CoV2 by RT-PCR in their NPS and saliva (collected simultaneously from the out-patients of the COVID clinic and in-patients of the COVID wards, VHS Hospital, Chennai, India) were selected retrospectively in an anonymous delinked manner. Ten saliva samples negative by RT-PCR (5 NPS-positive and 5 NPS-negative) were selected as controls. Twenty saliva samples from patients with NPS and saliva positivity by RT-PCR were selected as test samples. The presence of SARS-CoV2 antigen was tested using the Indian Council of Medical Research (ICMR) validated (for NPS samples) commercially available rapid lateral flow kit (SD Biosensor, Korea) [11]. Three hundred microliters of free-flowing saliva without any mucous material was mixed with the extraction buffer (provided in the kit). The test was performed as per the manufacturer’s instructions. The viral copy numbers in the NPS samples were extrapolated from the Ct (cycle threshold) values using the standard curve equation generated from commercially available SARS-CoV2 RNA standards of *N gene* and *RdRp gene* (Exact Diagnostics, USA). Median and interquartile ranges were calculated using Microsoft excel. The Mann-Whitney rank sum test and McNemar tests were performed using free online calculators.

All 10 RT-PCR negative saliva samples were negative for the presence of SARS-CoV2 antigens. Of the 20 RT-PCR positive saliva samples, 9 (45%) were positive. The sensitivity, specificity and concordance for the saliva antigen test were calculated against the saliva RT-PCR test. True positives were 9/20 (45%). True negatives were 10/10 (100%). Therefore, the sensitivity was 45%, specificity 100% and concordance 63%: the last being statistically significant (p=0.001; McNemar test). Antigen detection is generally less sensitive than RT-PCR. So, we compared the SARS-CoV2 copies in the NPS of the patients with a negative or positive antigen test. The median and interquartile range of the virus copies were markedly higher among the antigen positive patients (Figure 1) and this difference was statistically significant (p=0.01; Mann Whitney rank sum test).

**Figure 1:**
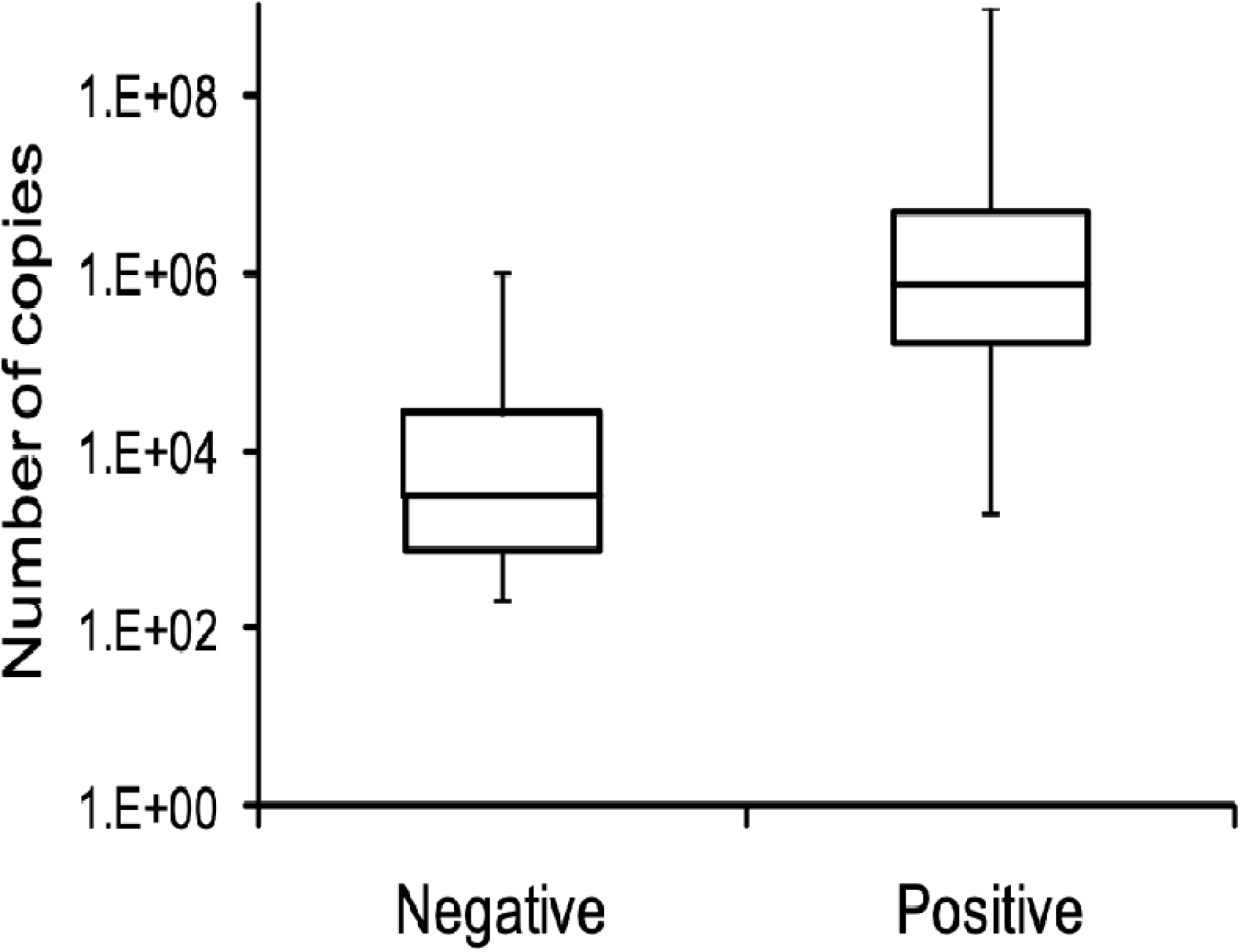
Patients with positive saliva antigen test have higher SARS-CoV2 copies in the NPS samples. X-axis denotes negative and positive categories of the saliva antigen test. Y-axis denotes the number of SARS-CoV2 copies. The interquartile range shows the 25-75% range of the virus copies in each category. The error bars depict the minimum and maximum copy numbers in each category.

Our data show that the point-of-care SARS-CoV2 antigen lateral flow test can detect the presence or absence of the virus in the saliva in 63% of the cases in a reliable manner. Additionally, the majority of the patients who were SARS-CoV2 antigen positive in their saliva samples had higher viral copies than those who were antigen negative. A similar result was shown by Nagura-Ikeda *et al*, although the sensitivity of their rapid antigen test was only 11.7% [12]. This difference in sensitivities could be attributed to the difference in testing methods. We have added free-flowing saliva directly to the extraction buffer. Nagura-Ikeda *et al* have dipped a cotton swab in the saliva sample, which was then dipped into the extraction buffer [12]. The cotton swab would have absorbed the liquid from the saliva with minimal absorption of virions and virus-infected cells thereby resulting is much less sensitivity. RT-PCR is a highly sensitive method that detects the presence of viral RNA by million-fold amplification. RNA from dead virus or unpackaged RNA will also be picked up by the RT-PCR. On the other hand, the antigen test has moderate sensitivity due to the lack of any kind of amplification and detects the translated proteins of the virus. Therefore, antigen positivity denotes abundance of proteins and in turn high copy numbers as shown in the figure. The limitation in the use of saliva samples is the requirement of a diligently collected free-flowing drooled sample without any sputum contamination, as the thick phlegm/mucous can compromise the free lateral flow of the sample across the chromatogram causing false negative results.

Overall, the point-of-care antigen test may be used for the detection of SARS-CoV2 in saliva samples for the rapid confirmation in symptomatic cases requiring urgent medical care. In post-treatment or post-quarantine people this test might be very useful to rule out risk of transmission, given that RT-PCR can remain positive for up to three months. Since this is a rapid, easy to use point-of-care test it can also be advocated to be used at home as a self-test. It would also be useful before invasive procedures, especially in dentistry, to assess the risk of transmission or in small remote medical centres where medical expertise to collect NPS samples and technical expertise for RT-PCR are often not available. The small group of antigen-negative people may be confirmed by collecting NPS samples and transporting to higher facilities for RT-PCR.

## Data Availability

All data pertinent to the manuscript has been uploaded with the manuscript files.

## Acknowledgements

This work was funded by intramural research funds of Chennai Dental Research Foundation, Chennai, India.

## Abbreviations

SARS-CoV2: severe acute respiratory syndrome – coronavirus 2
COVID-19: coronavirus disease 2019
RT-PCR: reverse transcriptase-polymerase chain reaction
NPS: nasopharyngeal swab
POC: point-of-care
RAT: rapid antigen test
ACE-2: angiotensin converting enzyme-2
ICMR: Indian Council of Medical Research

